# Assessment of breakthrough infections among post-vaccinated healthcare workers in a Tertiary Dental Hospital in New Delhi, India

**DOI:** 10.1101/2021.11.15.21266333

**Authors:** Deepamala Maurya, Arundeep Kaur, Farrukh Faraz, Shruti Tandon, Anju Rana, Shekhar Grover

**Affiliations:** Department of Periodontics, Maulana Azad Institute of Dental Sciences, New Delhi; Consultant-Dental Public Health, Mobile Dental Clinic Project, Maulana Azad Institute of Dental Sciences, New Delhi

**Keywords:** COVID-19, Healthcare workers, Infection, COVID-19 vaccine, Immunization

## Abstract

**Background:** COVID-19 vaccination in India has been rolled out on a national level, with healthcare workers (HCWs) becoming the first recipient of both Covishield and Covaxin. However, concerns over efficacy of vaccines have been much debated. This study highlights COVID-19 infections among vaccinated HCWs in a teaching dental hospital in Delhi, India.

**Methodology:** This cross-sectional survey was conducted using a pretested, validated, self-instituted questionnaire assessing COVID-19 like symptoms and/or confirmed infections among partially or fully vaccinated HCWs (all faculty, staff and students) of the institute from 16^th^ January to 31^st^ July 2021. The number of infections was also matched with hospital records.

**Results:** Out of 397 HCWs, 386 (97.2%) were vaccinated and 355 (89.4%) had received both doses. COVID-19 like symptoms appeared in 21 HCWs (5.4%) post any dose of vaccine. Symptomatic breakthrough infections >14 days after second dose occurred was seen in 16 HCWs (4.5%). Except one (required hospitalization), all other cases had mild infection. No significant difference was observed between Covishield and Covaxin. Most common symptom was fever and body ache.

**Conclusion:** The study identifies the possibility of breakthrough infections among vaccinated HCWs, and ensures the impact of vaccination in limiting disease severity. The findings suggest that COVID-19 preventive measures should be continued even among vaccinated individuals.

## Introduction

Coronavirus disease or COVID-19, caused by Severe Acute Respiratory Syndrome Coronavirus-2 (SARS-CoV-2), has gained worldwide attention since its origin in December 2019, and subsequent progression to a pandemic in March 2020. India has been one of its hotspots, reporting over 34 million cases and 4.5 lakh deaths by mid-October 2021^1^. Several measures have been implemented to control and combat this disease. Evidence suggests that COVID-19 associated disease severity, hospitalization, and death rates can be substantially reduced by Vaccination, in both clinical trial^2^ and real-world settings^3^.

The Government of India recognized the importance of COVID-19 vaccination, and permitted the clinical use of two vaccines from January 16, 2021, namely ChAdOx1 nCoV-19 (recombinant) vaccine [also called as ‘Covishield’ or Oxford-AstraZeneca Vaccine] and the whole viron inactivated vero cell vaccine or BBV152 [also called as ‘Covaxin’]^4^. The healthcare workers (HCWs) being the frontline warriors in the care of COVID-19 were initially offered the vaccination, and from May 2021 onwards, it was open to all individuals (>18 years of age) in India.

Although considered safe, there have been some concerns over efficacy and side effects of COVID-19 vaccines^5^. Since HCWs are at a greater risk of getting the infection, and might act as a source of further transmission as well, it is important to recognize the possibility of any COVID-19 breakthrough infection post-vaccination. Breakthrough infections, as suggested by CDC, are infections occurring >14 days after completing the primary series of vaccination^6^. A few cases of post-vaccination breakthrough infections have been reported among HCWs^7^. Hence, this study was conducted to assess COVID-19 infections post-vaccination among HCWs in a tertiary dental care hospital in New Delhi, India^8^.

## Methodology

The present study is a questionnaire-based, single-centre, cross-sectional survey assessing COVID-19 like symptoms [as listed by Center for Disease Control (CDC)^9^] and breakthrough infection in a tertiary dental care teaching hospital^7^. At the time of the survey, the institute had a total strength of 397 HCWs (including students and employees aged 18-60 years), out of which 386 (97.2%) had received COVID-19 vaccination. The hospital essentially maintains records of all vaccinations as well as COVID-19 infections of its HCWs.

All HCWs who were partially (only one dose) or fully (both doses received) vaccinated with either Covaxin or Covishield from 16^th^ Jan – 31^st^ July 2021 were included in the survey. A pre-tested and validated questionnaire was administered, which included socio-demographic questions and details of dates of vaccination, symptoms type & severity, home quarantine, hospitalization, and post-COVID complications (if any). Symptomatic HCWs were inquired if they had confirmed the infection by reverse transcriptase polymerase chain reaction (rt-PCR) or rapid antigen test (RAT). The impact of vaccination by antibody level testing was not done due to logistic reasons. The filled questionnaires were collected and matched with hospital records. Discrepancy, if any was rectified by telephonic confirmation of responses from the concerned HCW.

Ethical clearance was provided by the Institutional Ethical Committee, Maulana Azad Institute of Dental Sciences, Central Delhi, Delhi (India) (File No. EC/NEW/INST/2020/1207, dated 23/12/2020). The Institutional Ethical Committee approved the study. An informed consent was obtained from all participants prior to the distribution of questionnaires. The descriptive (frequency distribution) and inferential (chi-square test) analysis was conducted using SPSS v23.0 (IBM, New York, USA). For all analyses, p-value of <0.05 was considered statistically significant.

## Results

A summary of vaccination details and infection rates has been described in Table 1. A total of 97.2% HCWs were vaccinated, and 89.4% had received both the doses. Among the vaccinated, 62.4% received Covishield, 37% received Covaxin, and only 2 HCWs received some other vaccine (excluded from the final analysis). Only 2 HCWs (6.4%) had COVID-19 like symptoms after receiving the first dose, and 19 HCWs (5.4%) had symptoms after second dose. Nearly 4.5% HCWs experienced breakthrough infection. It was observed that dental faculty and students had a higher tendency towards receiving Covishield and the rest categories were administered more Covaxin (not depicted in table). The prevalence and onset of COVID-19 symptoms, as well as positivity rate were similar in both Covaxin and Covishield administered HCWs (p > 0.05) (Figure 1).

**Table 1:** Summary of vaccination and breakthrough infection among HCWs:

| Total HCWs | Vaccinated | Non-vaccinated | Doses taken |  | Type of Vaccine |  |  | COVID-19 symptoms |  | Breakthrough infection (COVID-19 test positive) |
| --- | --- | --- | --- | --- | --- | --- | --- | --- | --- | --- |
|  |  |  | One | Both | Covishield | Covaxin | Other | After 1<sup>st</sup> dose | After 2<sup>nd</sup> dose |  |
| 397 | 386 | 11 | 31 | 355 | 241 | 143 | 2 | 2 | 19 | 16 |

**Table 2:** COVID-19 symptoms and outcomes among infected HCWs:

| Symptoms | Overall prevalence |  | Outcome of COVID-19 infection |  | Overall prevalence |  |
| --- | --- | --- | --- | --- | --- | --- |
|  | N | % |  |  | N | % |
| Fever | 10 | 62.5 | Symptoms disappeared | Within 1 week | 10 | 62.5 |
| Cough | 9 | 56.3 |  | Within 2 weeks | 4 | 25.0 |
| Breathing Issues | 1 | 6.3 |  | Beyond 2 weeks | 2 | 12.5 |
| Fatigue | 1 | 6.3 | Quarantine period | None | 2 | 12.5 |
| Congestion | 1 | 6.3 |  | Upto 1 week | 2 | 12.5 |
| Body ache | 10 | 62.5 |  | Upto 2 weeks | 12 | 75.0 |
| Loss of taste & smell | 9 | 56.3 | Hospitalization required |  | 1 | 6.3 |
| Headache | 2 | 12.5 | Number of family members infected | None | 6 | 37.5 |
| Sore throat | 2 | 12.5 |  | Up to 2 | 5 | 31.3 |
| Diarrhea | 0 | 0.0 |  | More than 2 | 5 | 31.3 |

**Figure 1:**
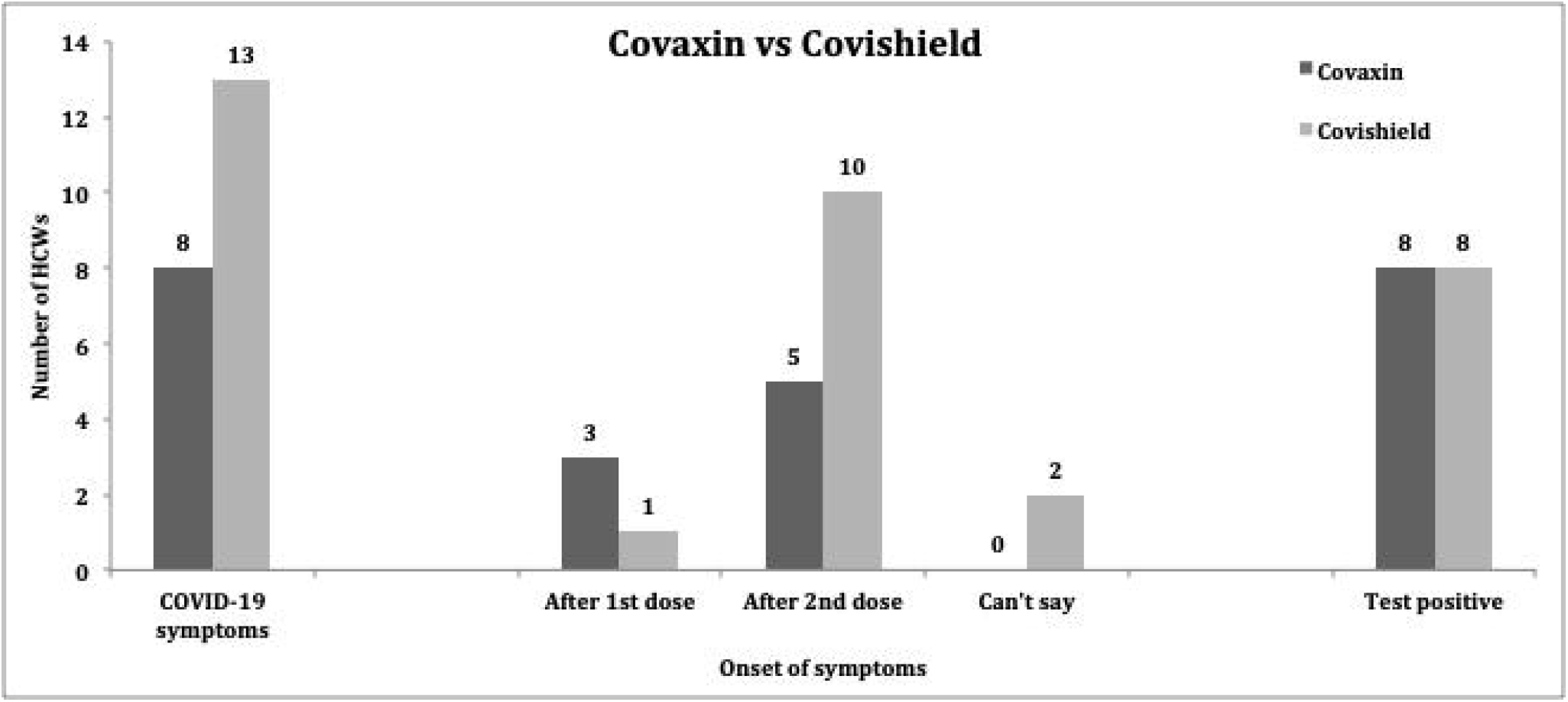
Comparison of Covaxin and Covishield regarding prevalence and onset of symptoms and positivity rate.

Table 3 presents details of various symptoms and outcome reported by infected HCWs. Fever (63%) and body ache (63%) were the most commonly reported symptoms. The prevalence of symptoms was similar in both Covaxin and Covishield administered HCWs. A majority of infected HCWs reported that their symptoms disappeared within 01 week (63%), and three-fourth of infected HCWs went for home quarantine up to 2 weeks (75%). Only 01 individual suffered serious symptoms and required hospitalization. Nearly 63% of infected HCWs passed on the infection to their family members.

## Discussion

Although health care workers (HCWs) from all specialties are at a high risk of exposure, the risk for dental professionals is theoretically more, owing to the proximity with the patients during oral examinations, substantiated by potential for direct exposure due to aerosol-generating procedures and use of high-vacuum suction^10^. Therefore, it becomes all the more pertinent to assess the real world effectiveness among this cadre of health care professionals. The present study showed incidence of breakthrough infections among HCWs to be 4.5% while 6.4% were having COVID-19 like symptoms. This is much lower than that reported from a similar study conducted by Tyagi et al (2021) among 123 HCWs in a chronic care facility in New Delhi^11^. This difference could be probably due to increased awareness levels and coherence to adoption of COVID-19 appropriate behavior among HCWs in this institution.

Every individual who was positive for COVID-19 had symptomatic appearance. Studies have reported a diverse percentage of infected HCWs developing symptoms. While study by Tyagi et al reported all HCWs with breakthrough infections to be symptomatic, a study by Teran et al among 75 skilled nursing care facilities in Chicago, Illinois reported that nearly two thirds (14 of 22; 64%) of persons with breakthrough infections were asymptomatic^12^. Another study, conducted among 1497 HCWs in a medical center in Israel revealed that only 33% of the HCWs with breakthrough infections were asymptomatic^13^. This also implies that vaccine does help in preventing symptomatic disease if not infection. These findings suggest that rigorous screening following even minor exposure helps in detecting asymptomatic infections and prevents further spread. No significant difference was observed between Covaxin and Covishield administrated individuals, indicating that both vaccine elicit a good immune response, consistent with other studies^14^.

The most common symptom presentation were fever and body ache among the infected HCWs in the present study. Other symptoms included cough, loss of taste and smell. Similar findings were reported in studies done elsewhere^11,13^. For 2 HCWs, symptom persisted for more than 2 weeks. The findings are similar to study by Tyagi et al^11^.

Among the 16 HCWs with breakthrough infections, six (37.5%) did not report any family member contracting the infection while 10 (62.5%) reported 2 or more family members getting infected with SARS CoV-2 in the present study. The updated systematic review and meta-analysis of 87 studies from 30 countries have reported secondary attack rate at household level to be 19%^15^. Higher rate in our study could be due to the fact that since our respondents were HCWs, they might have a higher viral load due to high risk of exposure, and hence the infectivity.

The cross-sectional nature of the study, single center assessment and small sample size limits the credibility of the study. Also, clinically relevant variables such as comorbities, age, gender, etc were not explored due to logistic reasons. However, this study strongly helps in identify the impact of vaccinations in real-world settings. It is comforting that the prevalence of breakthrough infections was less prevalent and mostly mild in severity, indicating that if not infection; vaccines have at least shielded the severe reaction from the disease. However, this may also create a possibility that there could be asymptomatic carriers among exposed HCWs.

Nevertheless, dental healthcare facilities being at the highest risk of infection are required to assess their HCWs for possible symptoms and spread of infection. Breakthrough infections thus should be focus of interest for healthcare institutions caring for COVID-19 patients and community dwelling individuals.

## Data Availability

All data produced in the present study are available upon reasonable request to the authors

